# COVID-19 Epidemic Dynamics and Population Projections from Early Days of Case Reporting in a 40 million population from Southern India

**DOI:** 10.1101/2020.04.17.20070292

**Authors:** Rashmi Pant, Lincoln P. Choudhury, Jammy G. Rajesh, Vijay V. Yeldandi

## Abstract

India reported its first COVID19 case on 30 January 2020. Since then the epidemic has taken different trajectories across different geographical locations in the country. This study explores the population aggregated trajectories of COVID19 susceptible, infected and recovered or dead cases in the south Indian state of Telangana with a population of approximately 40 million. Information on cases reported from March 2 to April 4 was collated from government records. The susceptible-infected-removed (SIR) model for the spread of an infectious disease was used. Transmission parameters were extracted from existing literature that has emerged over past weeks from other regions with similar population densities as Telangana. Optimisation algorithms were used to get basic reproduction rate for different phases of nonpharmaceutical interventions rolled by the government. Peak accumulation is projected towards end of July with 36% of the population being infected by August 2020 if the population lockdown or social distancing mechanism is not continued. The number of deaths assuming no intervention is projected to be 488000 (95% CI: (329400, 646600)). A draconian enforcement of population lockdown combined with hand and face hygiene adherence would reduce the transmission by at least 99.7% whereas partial social distancing and hygiene would reduce it by 51.2%. Transmission parameters reported should be interpreted with caution as they are population aggregated and do not consider unique characteristics of susceptibility among micro-clusters and vulnerable individuals. More data will need to be collected to optimize transmission parameters and evaluate the full complexity, to simulate real world scenarios in the models.

## Introduction

The announcement of the novel corona Virus (COVID-19 or SARS-CoV-2) as pandemic was made on January 30^th^ 2020 [1]. The first case of COVID-19 was detected in India on January 30^th^, 2020. As of 30^th^ March 2020, more than 1250 cases had been identified in India, with 32 deaths and 102 cases have been discharged after treatment [2]. Many key aspects about the disease dynamics are not known. To improve the understanding about the virus many researchers continue to contribute through peer review journals, blogs, reports and social media platforms [3]. One of the key endeavours among these knowledge products is the quest to quantify the burden of disease through the use of mathematical modelling [4,5,6] so that public health systems can prepare for emergency response.

India is a geographically, climatically and culturally diverse country with nearly 1.3 billion population [7]. The population density not only differs from urban to rural areas but also from state to state with Delhi having more than 11,000 people per square kilometre, while Arunachal Pradesh has only 17 people per square kilometre. The country has 53 cities which have more than a million population with a minimum density of 400 persons per square kilometre, according to the census of 2011. The country has 137 airports, including 23 international airports handing more than 6 million international and 20 million domestic passengers every month. The above information indicates the diversity of population distribution that can influence the spread of an infectious disease like COVID-19 and the possibility of import via international passenger influx [8]. Though there have been recent publications on the estimated size of the epidemic in India, at times the methodology or the tools to replicate the same is not available in public domain limiting the access and validation of such tools [5,6] This paper describes a simple mathematical model to understand COVID-19 epidemic using observed data and provides a free tool that is available for anyone including the states, local governance system managers to download and use it to have a better understanding of the scale and progress. We present model projections for the Telangana state in southern India which has a population of around 39.64 million people and the sixth busiest international airport in India [9].

## Materials and Methods

There are several mathematical models available and used for the different diseases including COVID-19 [10,11,12]. We used the well-known susceptible-infectious-recovered (SIR) Model for infectious diseases [12]. This is a simplistic yet effective *compartmental model* where individuals in a target population start from the compartment of “Susceptible” and upon infection, individuals move to the “Infected” compartment and subsequently they move to the recovered or removed compartment based on disease outcome. An inherent assumption of the simple SIR model is that every individual in the compartment has similar characteristics. The limited information on COVID-19 epidemic dynamics informs us that the virus behaves like the Severe Acute Respiratory Syndrome (SARS) epidemic family. The SARS infection was associated with a high level of immunity after infection [13]. We assumed that COVID-19 creates similar immunity in the human body reducing the chance of reinfection. Also, there is no documented case of reinfection of COVID-19 in the current epidemic in the Republic of China or elsewhere. Thus, we assumed that all those who are infected will be “removed” from the pool of susceptible either due to recovery or death. Because of the short nature of the epidemic elsewhere we assumed that the epidemic is not affected by larger population-level vital dynamics i.e. births, migration etc.

The SIR model is represented mathematically by a series of differential equations given below.

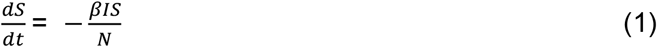

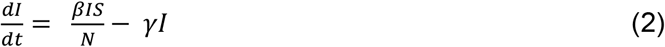

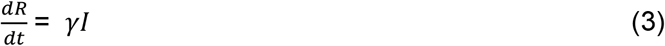

Where S= Susceptible Population, I= Infected Population, R= Removed (Recovered or Died). S+I+R=N

For the purpose of our analyses, time (t) is the time since the first COVID19 case as reported in Telangana state. For the purposes of this analysis, we have assumed a start date of 2 March, 2020 when the state of Telangana diagnosed its first case.

### Choice of parameter and optimisation

We first calculated projections using transmission parameters reported by other geographical locations with a similar population density [Wang H et al (14,15]. This will allow us to confirm population level disease dynamics of COVID19. We the used least squares optimisation to calculate transmission parameters based on actual reported data from the state. The transmission parameter (*β*) from the S to the I compartment, defined as the average number of contacts per person per time was determined to be in the range 0.05-0.17. The transmission parameter from I to R compartment, (*γ*) known as recovery rate was fixed at 1/18 (=0.056). This corresponds to a recovery period of approximately 18 days. The model parameters were estimated using the sum of squares method and optimised using Limited-memory BFGS method [16]. The estimated incidence and the reported cases were re-checked visually for their fit.

We assumed a per person contact rate of 40 individuals and an initial infection probability of 10% to arrive at estimated number of initial infected cases as 4. The gamma parameter for our SIR Model was fixed at 1/18 [14] as it agrees with observed data where the first five reported recoveries were after 16-20 days of isolation. We advise a note of caution here as the reported infected cases during the initial days of the outbreak may be an underestimate of the burden of disease due to limited testing in the country.

The *β* and *γ* values were used to calculate the basic reproduction number, R_0_, which measures the transmissibility of a virus, representing the average number of new infections generated by each infected person. R_0_ > 1 indicates that the outbreak will continue to yield increasing number of infections unless effective control measures are implemented, while R_0_ < 1 indicates that the number of new cases decreases over time and, eventually, the outbreak will end. Thus, R_0_ is a time-varying measure whose periodic assessment during an epidemic informs policymakers on the need for and effectiveness of interventions. We obtained projections for infections and mortality by calculating R_0_ by least squares optimisation and also based on a range of *β* values from 0.07 to 0.17 [14] for three different phases of nonpharmaceutical intervention (NPI) launched by policy makers in the state of Telangana. These phases are:

A. The R_0_ arrived using the optimisation of the observed cases in Telangana state from 2 March 2020 to 4 April 2030
B. 2 March 2020 to 15 March 2020 (no intervention and limited face and hand hygiene messaging).
C. 16 March 2020 to 25 March 2020-voluntary social distancing (work-from-home and “Janta curfew” advisory [17]) and setting up of quarantine and isolation beds.
D. 26 March 2020 to 14 April 2020-population lockdown announced including closure of international airports and cargo ports
E. 15 April 2020 onward post lockdown with partial resuming of population movement

The R_0_ for these scenarios is calculated to be (A) 1.38; (B) 3; (C) 2.6; (D) 1.9 and (E) 2.6. To obtain projections for mortality due to COVID19, we fit a simple moving average [18] of order 3 to the case fatality rate (CFR) observed over the 34-day period.

All analysis was done using R-software (version3.3.3). The daily case report data from Telangana used for preparing figures 1,2 and 3 is available in supplementary table 1.

**Figure 1:**
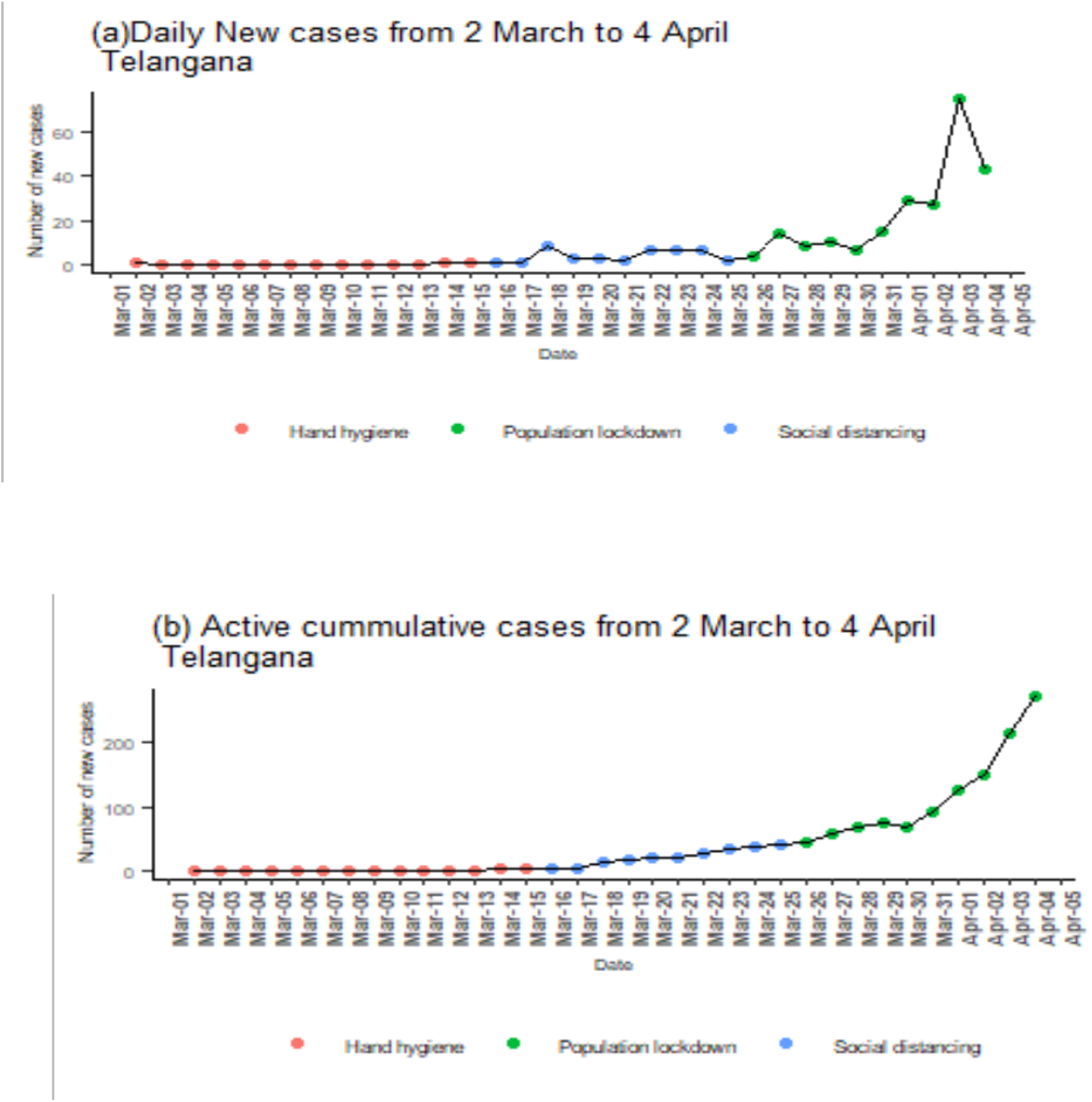
(a) Daily new COVID19 positive cases in Telangana from 2 March 2020 to 4 April 2020 by three different phases of nonpharmaceutical intervention (b) Active cumulative COVID19 positive cases in Telangana from 2 March 2020 to 4 April 2020 by three different phases of nonpharmaceutical intervention

**Figure 2:**
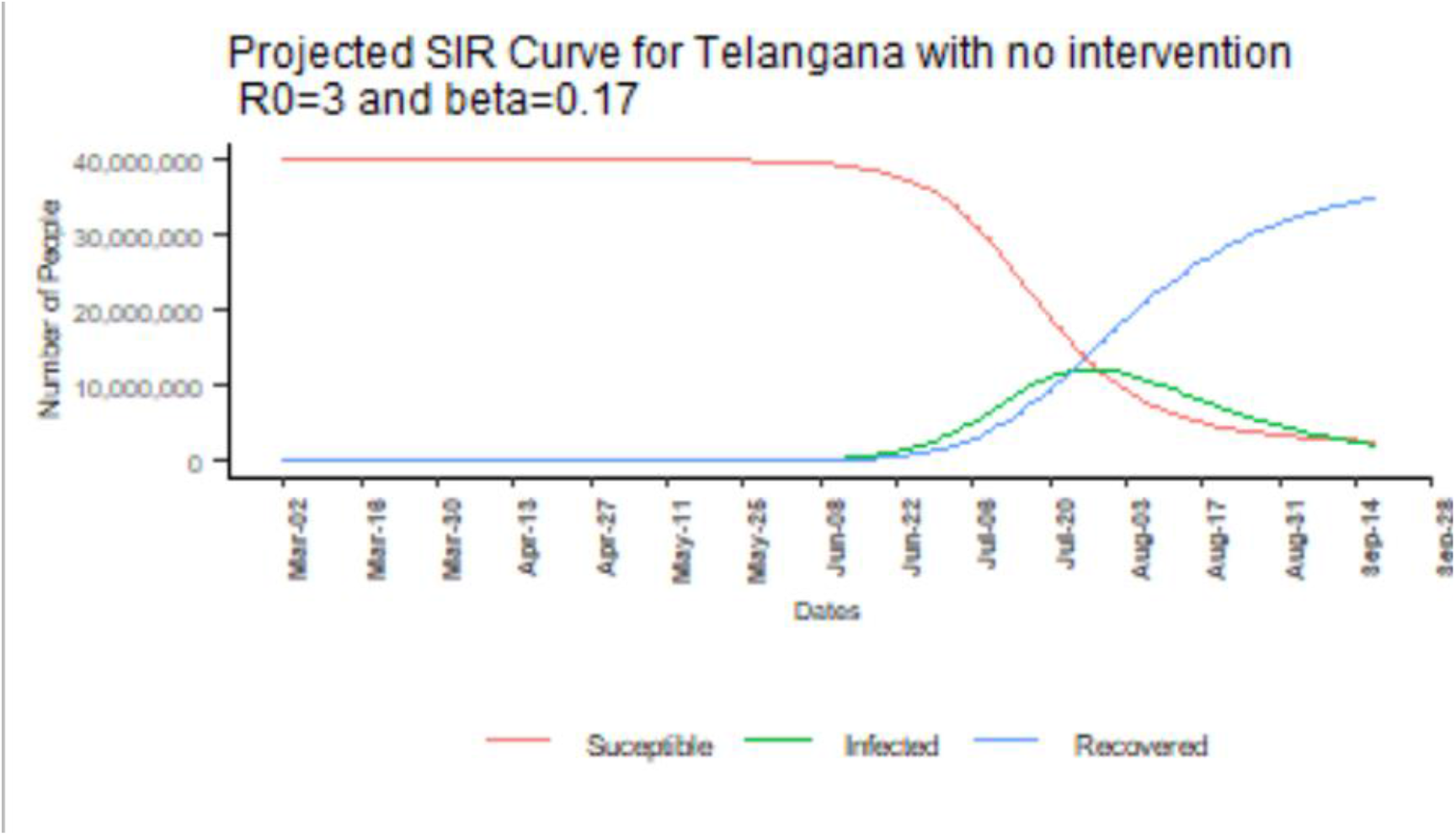
Projections of COVID19 spread for Telangana from SIR Model

**Figure 3:**
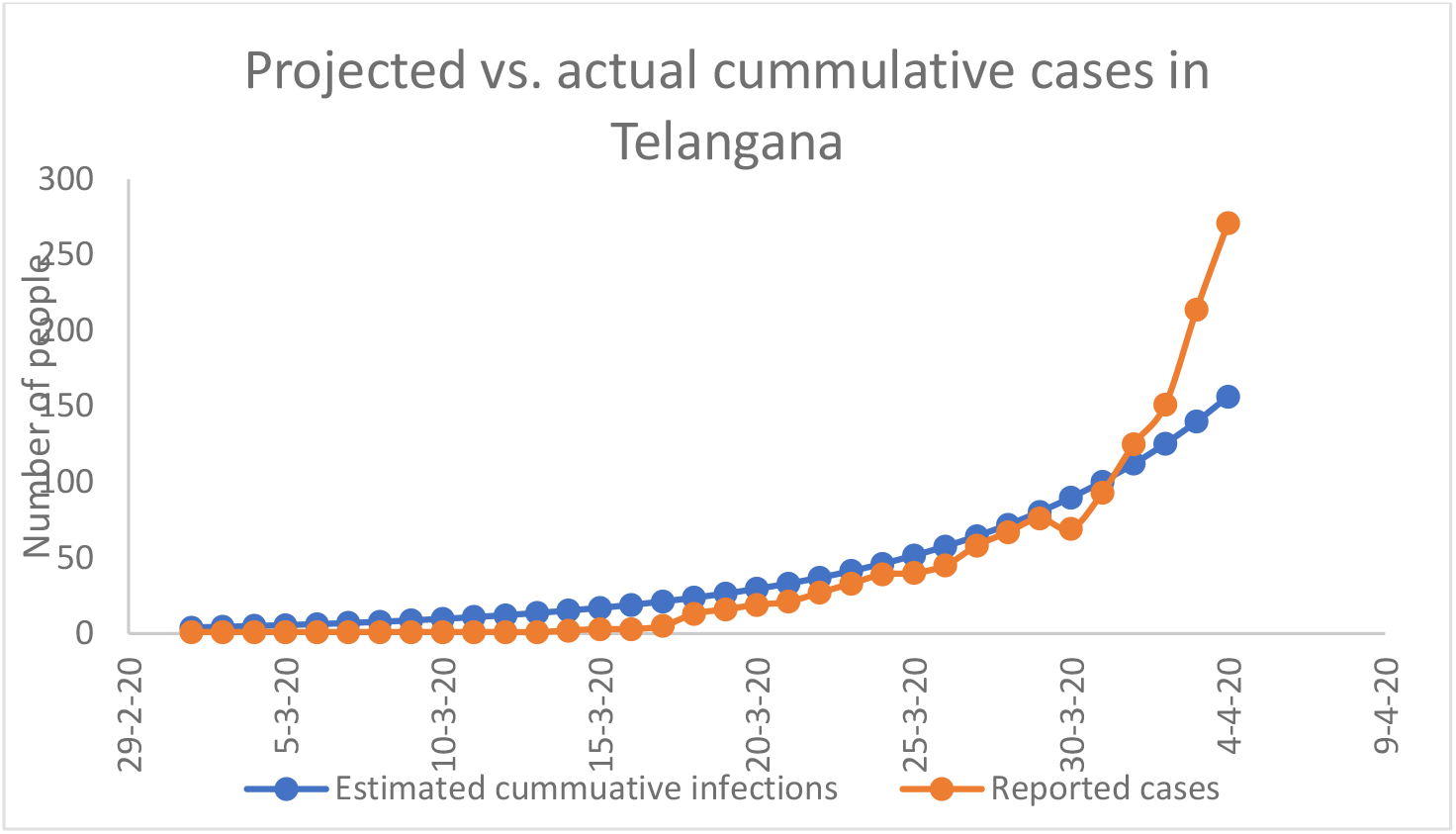
Fitting SIR Model to reported cumulative COVID19 cases in Telangana (2 March 2020 to 4 April 2020)

## Results

Based on available data as of 4 April 2020(Ministry of Health and Family Welfare, www.covid19india.org), of the 272 people who tested positive, 33(12.13%) recovered and 11(4.0%) died. The figures below show the incident cases and cumulative cases after removal of recovered and died individuals during each phase of the NPI rolled out by the government.

Projection of the spread of the infection in the Telangana population using the SIR model are presented in figure 2. The model (Figure 2) shows that in the absence of stringent interventions (R0=3) infections would peak towards the end of July and first week of August. An estimated number 11,910,208 individuals (36% of the population) of Telangana would have COVID19 infection during that time. The WHO estimates [19] that in India, 80% of the infections are currently asymptomatic or mild,15% are severe enough to require hospitalisation and 5% need critical care (ICU with ventilator). By extension, this would translate to a public health requirement of at least 2,382,042 hospital beds for sever or critical patients at the projected peak of infection by first week of August 2020. The state of Telangana currently has 277,850 hospital beds.

Figure 3 shows that the R_0_ parameter value of 3 fits the initial observed cases quite well. The population lockdown started on March 25, so we can see the accrued effect of no intervention in the gradual increase in reported cases during March 26 to April 4.

A sequential simulation of cumulative infection projections with distribution of the infected population by different phases of NPI initiated by the government of Telangana, is presented in figure 4.

**Figure 4:**
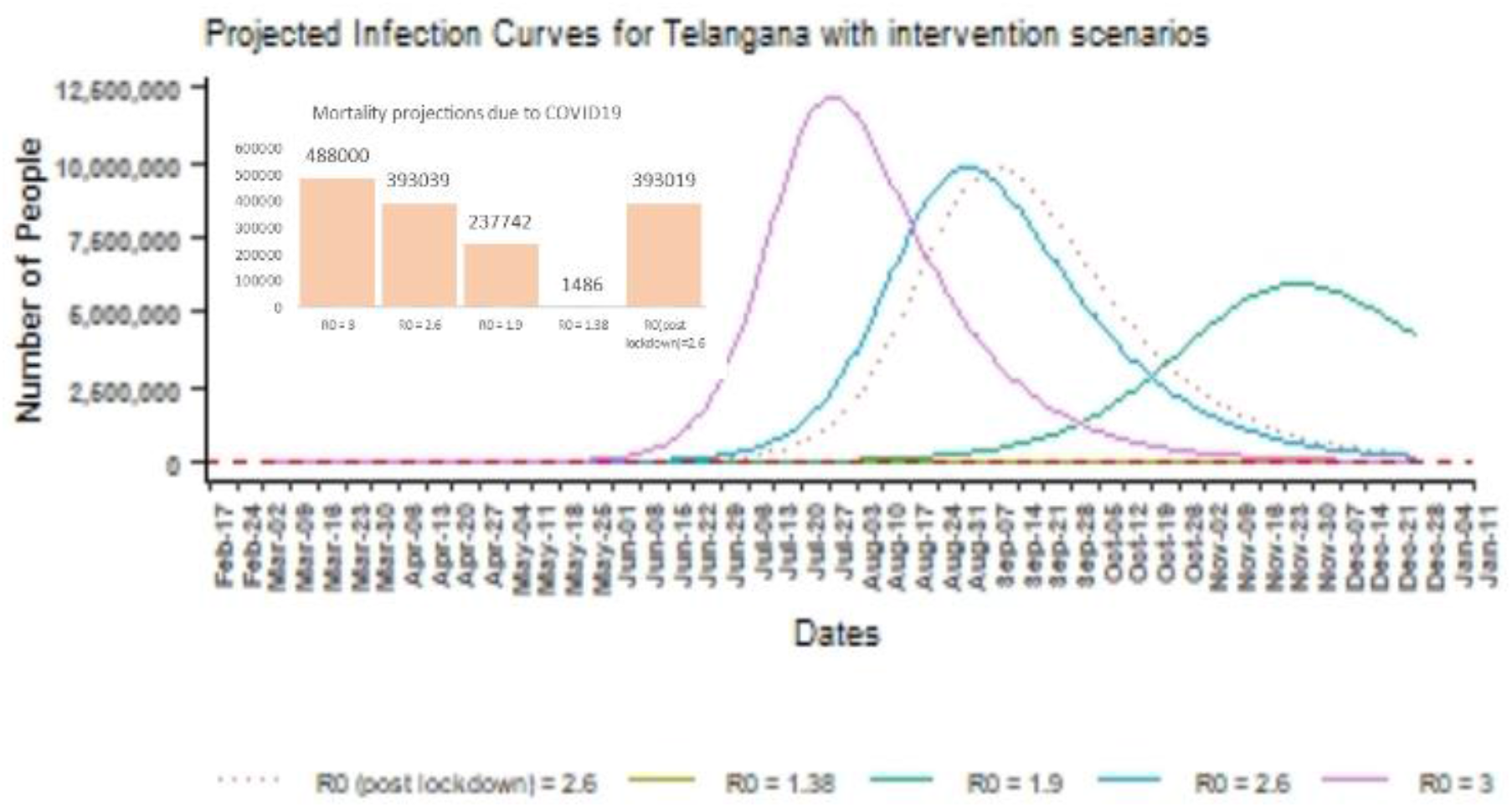
Projections of infection spread and mortality at peak with phased nonpharmaceutical Intervention scenarios from 2 March 2020 to 27 December 2020.

Figure 4 also shows that with measures such as face and hand hygiene and public advisories on adherence to social distancing (NPI-C)), the peak cumulative number of infections can be reduced by only 51.2% approximately (peak infections reduce from 11,910,208 to 5943550). With a complete population lockdown continuing beyond the stipulated 21 days, the peak infections reduce drastically by 99% indicating a dramatic and significant “flattening of the curve” [20]. Using the method of least squares to optimise the raw sum of squares using the L-BFGS-B method, the optimal values for β was 0.58 and γ=0.42 which gives an R_0_ of 1.38 for the initial 34 days of the observation period. Assuming this to represent the trend of cases in the state, the number of active cases at peak infection would be 37,157 and would yield death equal to 1486 (95% CI=(1003,1969)).

A simple moving average of order 3 was superimposed on the case fatality rate (CFR) obtained during the observation period of March 2 to April 4. This moving average series was used to forecast the CFR over a 60-day period. The average CFR was 4% with 95% confidence intervals (2.7%,5.3%). So even at the peak of infection in the voluntary social distancing and hand hygiene adherence phase, we could expect at least 2,37,740 fatalities with 95% CI (160,476, 315,008).

## Discussion

The epidemic curve presented in the different scenarios give a range of the burden of the disease and the scenario of optimisation with the state level data gives a very optimistic scenario. Although the range of scenarios with and without interventions gives us a spread of projections, we however feel that the no interventions scenario may be close to the actual scenario till the second week of April as accumulated cases will reflect undiagnosed infections and unreported deaths in the community. This model considers data till ten days after the shutdown announcement by Government of India on 25 March 2020, which is closer to the incubation period of the disease, indicating that the infections detected in last ten days are not influenced by the shutdown. However, there are other measures like a ban on international travel, campaign on handwashing etc. that was ongoing for almost four weeks, thus the influence of the same on the epidemic progression must be factored in. After the peak around 120 days from the first detected case, the epidemic is expected to show a decline in numbers and be on the downward slope of the curve. Similar estimates are published for the duration of the epidemic in the United kingdom [21]. Assuming the most optimistic outcome of the NPI as presently envisaged, at the peak, the epidemic will lead to proven 37,157 infections in the state of Telangana. However, this information needs to be contextualised with the natural history of the disease. the epidemic dynamics known to date and quality of reporting. COVID 19 is known to have asymptomatic infection among 17% of the infected population [22]. Based on data WHO reported [19], nearly 80 per cent are expected to have a mild or asymptomatic infection, which may manifest as a mild upper respiratory infection resembling mild common “flu” with severe cases needing critical care/ventilation of around 5%. Some authors have suggested that the actual mortality of COVID-19 may be much lower than what is reported and possible range between 0.25 per cent to 3 per cent, with their opinion favouring the lower estimates [23–27].

In a national COVID model [24], the authors suggest two types of containment strategy i.e. Port of entry and, (ii) mitigation – within-country connectivity. One of the arguments for the epidemic response was to have a robust screening at ports of entry and contact tracing program. Our preliminary model for Telangana state does not incorporate strategy (i). The capital city of Hyderabad has one international airport with total traffic of more than 20 million in the year 2018-19, including 4 million international travellers. The city has good railway and road connectivity to other important metro cities like the national capital of Delhi, Mumbai, Bengaluru and Chennai. Thus, the city is one of the high-risk areas for COVID-19 transmission. Other than being in the time of a highly transmissible virus-like COVID-19, the city has a high population density with 18,172 persons per Sq. Km [28]. One of the limitations of this work is the lack of discrimination between urban and rural areas. This was deliberate as at the time of collating data, reports stratified by level of urbanization were not very reliable. However, the possibility of the infection/epidemic already moving beyond the city perimeter to other districts or rural parts of the state cannot be ignored.

It is well known that with the availability of new information, in recent years, the country changed epidemic estimates for other epidemics like HIV and TB [29,30]. Also historical experiences from earlier outbreaks [31 should be combined with new estimates to inform effective interventions. Any scientific estimation needs robust local data. COVID-19 is new, and as one moves in time, more evidence will be available for better estimations.

The authors would like to emphasise that, these are population level projections. The inherent assumptions will not address micro clusters such as health workers, the modelling does not adjust for vulnerable groups and loci that may be high risk locations such as hospitals. At the present time more data is needed to clearly understand the differential transmission dynamics in special groups. The model does not have the ability to project precisely what may happen after the lockdown is lifted, previous experience with the 1918 Influenza pandemic [31] suggests that many different possibilities exist. Measures such as lockdown are considered as drastic public health measures with their long-term benefits unclear but may also have varied impact on the society [32–34].

## Conclusion

The outputs of this model show an expected population-level decline in the burden of reported infections/disease over time. The input data is influenced by the series of measures implemented locally by the authorities, thus its influence over the trajectory of the epidemic cannot be overlooked. As policymakers walk the tightrope of initiating public health interventions to contain the COVID19 epidemic, more granular analyses will be needed, especially in a country as socially and geographically diverse as India.

## Data Availability

The authors used an open-source program (RStudio-version3.6.3) that is widely used and leaving the codes to be accessed by other researchers on Github (https://github.com/). All the data used in the analysis will be available in the supplementary material.

## Data Availability

All data used in the manuscript is available in the public domain. Daily case summary reports of COVID-19 for the state of Telangana were obtained at https://covid19.telangana.gov.in/

https://figshare.com/articles/Telangana_Lincoln_csv/12151539

https://www.covid19india.org/

## Conflicts of Interest

The authors declare that they have no conflict of interest to report.

## Funding Statement

This study was funded by SHARE-INDIA internal research funds.

## Supplementary Materials

The data used in the analyses is available in Supplementary Table S1.

## Notes

### Competing Interest Statement

The authors have declared no competing interest.

### Funding Statement

No external funding received.

